# INVESTIGATING HEALTH PROFESSIONALS’ UNDERSTANDING AND RISK PERCEPTION OF THE EFFECT OF CLIMATE CHANGE ON HEALTH. A CROSS-SECTIONAL STUDY OF HEALTH PROFESSIONALS AT THE SDA HOSPITAL, REGIONAL HOSPITAL AND MUNICIPAL HOSPITAL-SUNYANI, GHANA

**DOI:** 10.1101/2023.05.24.23290473

**Authors:** Antwi Joseph Barimah, Mansurat Abdul-Ganiyu, Mohammed Mohammed Ibrahim, Solomon Saka Allotey, Rebecca Dorcas Commey, Angelo Guapem Osei-Tutu, Yaw Boakye Nketiah, Bernard Opoku Amoah, Larry Agyemang, James Dumba, Helina Gyamea

**Affiliations:** College of Health, Yamfo; National Ambulance Service; Travellers Trinity Clinic

**Author notes:** Corresponding Author Antwi Joseph Barimah.

**Keywords:** Climate Change, Risk Perception, Global warming, mitigation, health professionals

## Abstract

The study investigated health professionals’ understanding and risk perception of climate change in the Sunyani Municipality by focusing on health professional’s knowledge on climate change, examining health professional’s risk perception of climate change on health and identifying the co-benefits of climate change mitigation. This quantitative oriented cross-sectional study randomly selected 400 health professionals across the Regional Hospital, SDA Hospital and Municipal Hospital as respondents. Quantitative data was analyzed using SPSS Version 25. From the study, the results indicate that Health professionals are fully aware of the concept of climate change. Health professionals do not have knowledge pertaining to the scientific aspects of climate change. There was a statistical relationship between respondent’s perception that climate change can lead to death (P<0.001) and their awareness of the risk of climate change impact on health. There was a general likelihood of an increase in malaria (Mean=2.98), Dengue fever (Mean=3.16), Cholera (Mean= 3.18), schistosomiasis (Mean=3.27), Meningococcal meningitis (Mean=3.85) and Influenza (Mean=3.73) due to climate change. These actions positively affect health and climate and they include: Giving up red meat (Mean=3.21), Walking and cycling instead of using cars (Mean=3.27), Reducing rural-urban migration (Mean=3.46), Reducing air pollution from emission of fossil fuel (Mean=3.63). A majority of 65% of respondents agreed to the incorporation of climate change related course work into nursing/medical school curricula as a policy to mitigate climate change. The study concludes that health professionals are fully aware about climate change but lack a thorough understanding of the scientific aspects of climate change. The general risk perception of health professionals towards climate change impact on health was high. Climate change mitigation is beneficial to human populations.

## Introduction

Global warming is unambiguously apparent, as objective scientific observations are evidenced by surging sea levels globally, the continuous melting of snow, increment in ocean temperatures and the average warming of global air.^8^

Lately, climate change has garnered enormous attention in the global environmental discourse. This huge premium attached to climate change as a result of its devastating impact it visits on the living conditions of human populations. The concept of climate change is at the heart of developmental discussions globally. Apparently, low income countries especially countries in African Sub-region have shown great concern about climate change due to their vulnerability to the negative repercussions associated with climate considering their economic situations.^1^ Literature have effectively shown the negative impact climate and its attendant weather pose on the key determinants of human health such as water for drinking, food for energy and air to sustain life on earth. Undoubtedly, climate change and weather precipitates the transmission chain of infectious diseases among populations, drive storms and trigger heat waves which also pose an existential threat to human lives.^12^ The continuous emission greenhouse gases and carbon dioxide due to agricultural and industrial activities makes climate change a dangerous phenomenon that can alter the health of the human race.^12^

Climate change continues to visit mayhem on the human race with its protracted flooding, horrendous droughts, scary heat waves, rising ocean levels, distorted rainfall patterns and abnormal temperatures. Sadly, human beings are the most affected by either directly or indirectly as flooding, altered air quality, food shortage and poor nutrition due to drought, rising ocean levels, excessive temperatures etc all can lead to suffering, disabilities and death in most circumstances.^22^ Evidence from literature has shown that developing and developed countries alike are not spared by the devastating impact of climate change as the severe forms of drought have occurred in Africa and South Asia, monstrous tidal surges and devastating cyclones have occurred in Europe and the United States (e.g. Hurricane Katrina). However, a thorough comparative analysis between developed and developing countries reveal that the developing world are highly predisposed and more vulnerable to the adverse effects of climate variability and face more threats to the health of their people especially when such countries do not have the financial muscle to deal with the health risks posed by climate change.^20^

It is worth mentioning that in most developing economies, the smallest increment in global warming greatly affect agricultural production since crops are sensitive to extreme temperatures, this situation affects the economies of such agrarian developing economies. The corresponding health impact of malnutrition, dengue fever, malaria, diarrheal diseases continue to kill millions in such developing countries especially sub-Saharan Africa.^4^ The preceding revelations were further highlighted in the UCL Lancet Commission report, which opined that the most severe form of disability adjusted life years (DALYs) of 34% occasioned by climate change is projected to affect sub-Saharan Africa. It is worthy of note that Since sub-Saharan Africa only represents 11% of the global population, this situation reflects a looming health threat due to the adverse repercussions of climate variability which threatens the collective survival of sub-Saharan Africans.^4^

This therefore necessitated the investigation of health professionals’ understanding of the effects of climate change on health in the Sunyani Municipality of Ghana by focusing on health professional’s knowledge on climate change, examining health professional’s risk perception of climate change on health and identifying the co-benefits of climate change mitigation.

## Methods

### Study Design

A descriptive cross-sectional design was adopted for this study. The study employed descriptive study in order to get a clear picture of health professional understanding and risk perception of the effects of climate change on health. Thus, data was collected from the targeted health professionals over a short period of time (from March to May 2022).

### Study Population

The targeted population for the study was health professionals (specifically Doctors & Nurses) who were working in the Sunyani municipality. The three health facilities selected were SDA hospital, Regional hospital and Municipal hospital. The study employed SDA hospital, regional hospital and Municipal hospital because they are the biggest health facilities within the Sunyani Municipality. Hence, researchers randomly selected doctors and nurses across the three health facilities.

### Sample Size

The sample size was calculated using a 95% confidence interval, at a 5% margin of error. Yamane, Taro (1967) formula for calculating sample size, was used to compute the sample size. 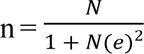 where; **n** is the Sample size, **N** Signifies the population of Sunyani municipality, and **e** = The desired level of precision (margin of error).

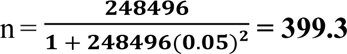

The sample size was rounded up to **400**.

### Data Collection

The study relied solely on primary data that was collected using a structured questionnaire. The questions encompassed both open-ended and close ended questions. Data collection techniques employed included one on one interviews using the structured questionnaire. Quantitative data was thus obtained from respondents using questionnaires. The data collection was done using English language throughout the entire data collection process. Data after have been collected were cleaned to ensure validity before data entries were made.

### Data Analysis

Structured questionnaires were used to solicit quantitative data. Data were cleaned to ensure validity before data entries were made into **SPSS** (statistical package for the social sciences) version **25** for rigorous statistical analysis. Data was presented using frequency as tables and charts as presented under the results section. Correlative statistics using Pearson chi-square test and binary logistic regression was used to show strengths of association between independent and dependent variables where appropriate.

### Ethical Considerations

Since the study involved the use of human subjects, ethical approval was sought from Ghana Health Service Ethical Review Committee who rigorously scrutinized the protocols before finally granting an ethical backing to the study. Informed permission was further sought from the Municipal Director of Health Services as well as management of the three selected health facilities used for the study. Permission was also sought from the staff before the study commenced. The purpose of the study and its attendant possible risk and benefits were thoroughly explained to research participants and they were further assured of utmost confidentiality of the information they provided and the fact that it would only be used solely for the purpose of the study. Hence, formal written consent was obtained from participants prior to data collection.

## Results

**From the table 1** above, it shows that, a little above half of health professionals (50.5%) were female and 49.5% were male, 29.5% were between the ages of 35-44, 18% were within the youngest category of 15-24 and 5.25% were between 55-60 years. On ranking, majority (92.5%) were nurses and 7.5% were doctors. Out of 100%, 72% were Christians, 20% were Muslims and 5% were from the African Traditional Religion. Majority of the respondents (99%) studied in Ghana and 1% studied outside Ghana.

**Table 1:**
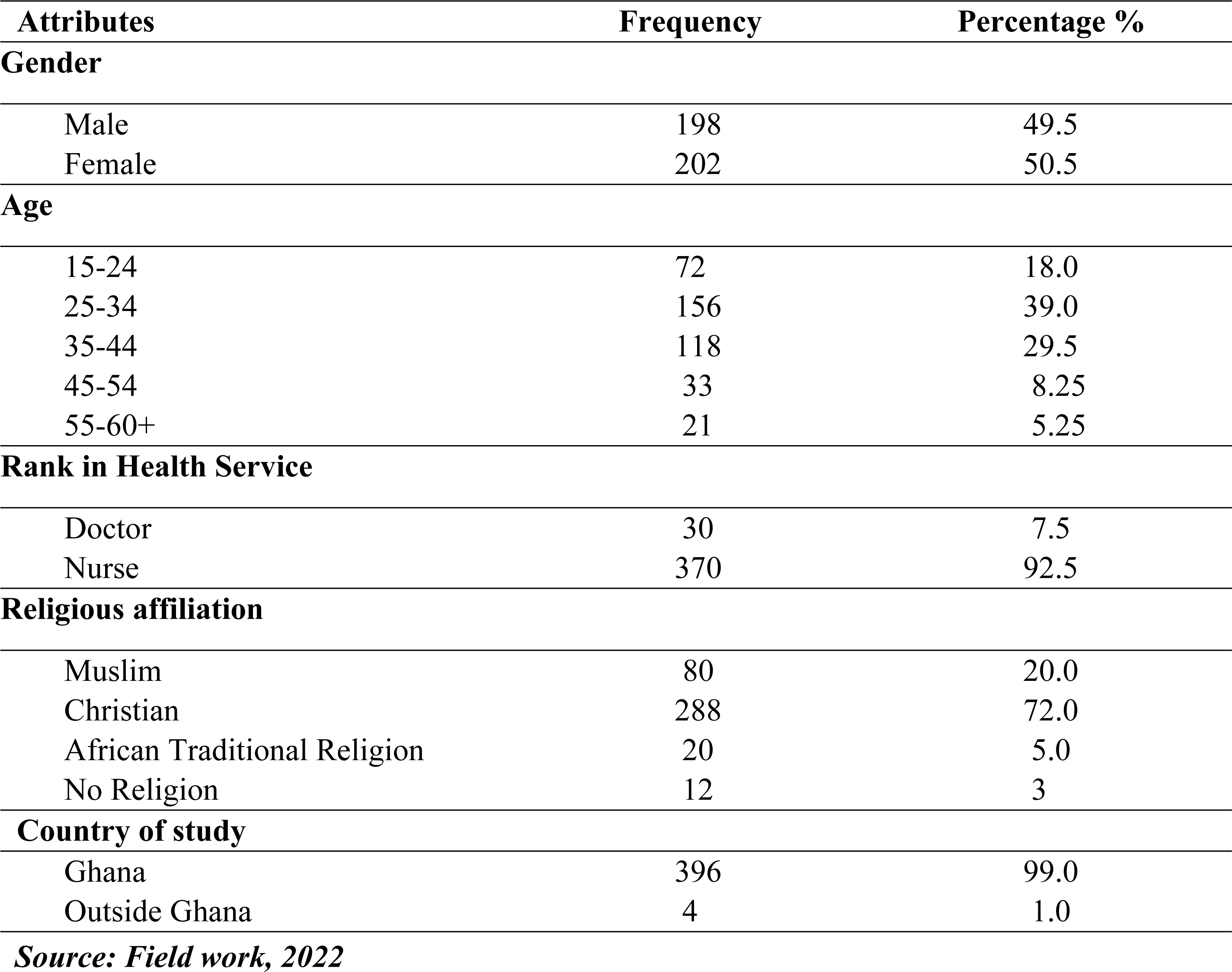
Background Attributes of respondents (N=400)

On health professional’s knowledge on climate change, all respondents (100%) had awareness on the concept of climate change. Majority of the respondents (87%) agreed that human activity is responsible for climate change, 75% noted natural variability as responsible for climate change. With respect to the extent of human and natural activity on climate change, more than half (65%) indicated both human and natural activity on climate change, 17% mainly claimed it on natural variability, 13% on mainly human activity and 5% on solely natural climate variability. As to whether climate change awareness campaigns are carried out within Sunyani Municipality, 39% indicated “yes”, 38% indicated “no” and 23% didn’t know anything about these awareness campaign. Out of 100%, more than half (70%) emphasized both developed and developing country as vulnerable to the effects of climate change, 11% indicated developing countries and 7% stated developed countries.

**Figure 1** shows respondents’ sources of information on climate change; 19% of the respondents get information on climate change from the social media, 17% from the radio, 16% from television stations, 15% from weather forecast, 14% from the internet, 12% from newspapers and 9% from friends and family. (Find attached)

**Figure 1.**
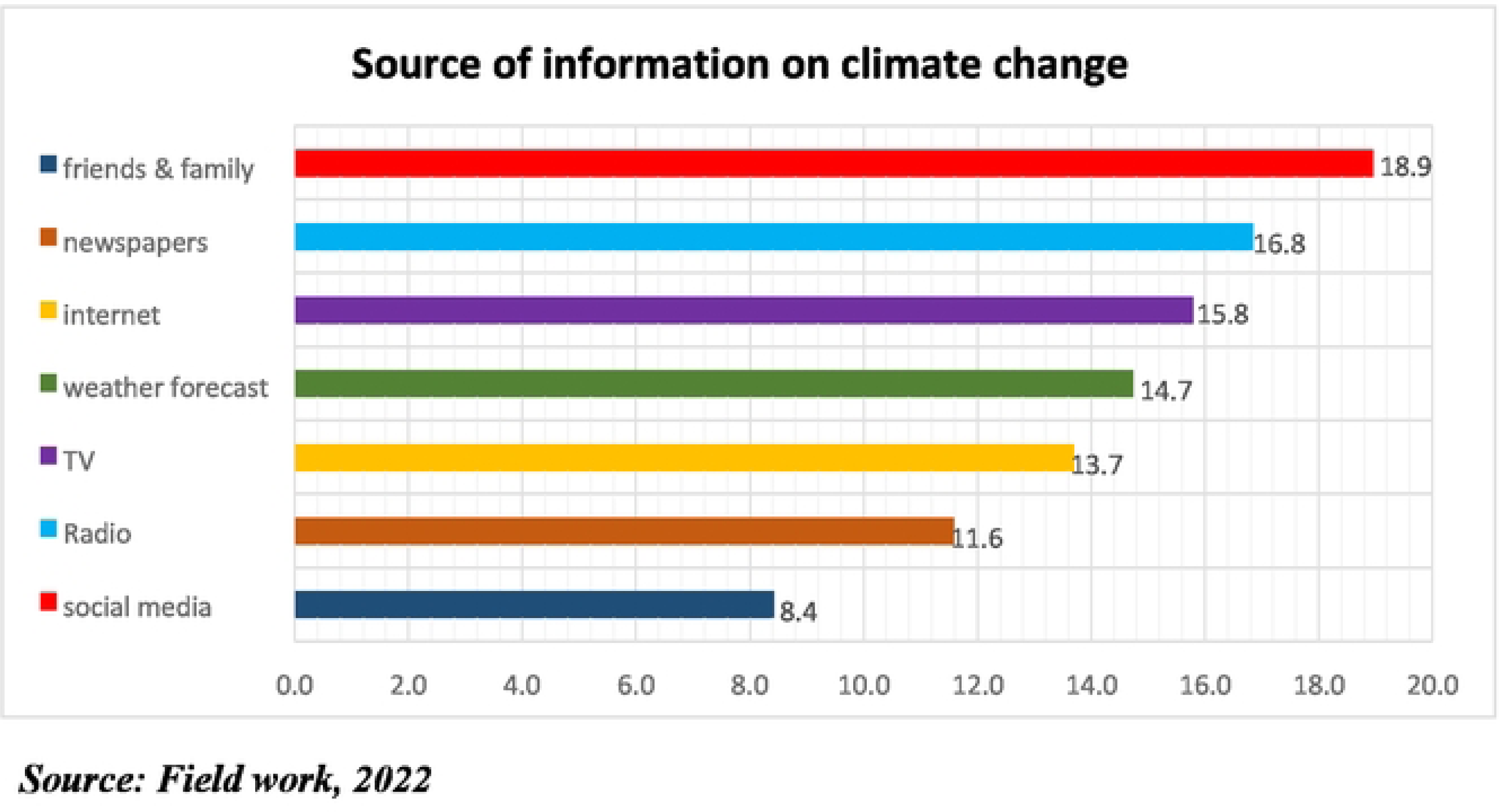
Source of information on climate change.

**Figure 2** displays the effects of climate change, 36% stated rising temperature, sea level rise, melting glaciers and drought as the possible effects of climate change, 20% indicated raising temperature, 16% noted rising temperature and flooding, 10% indicated rising temperature, melting glaciers, flooding and drought. (Find attached)

**Figure 2:**
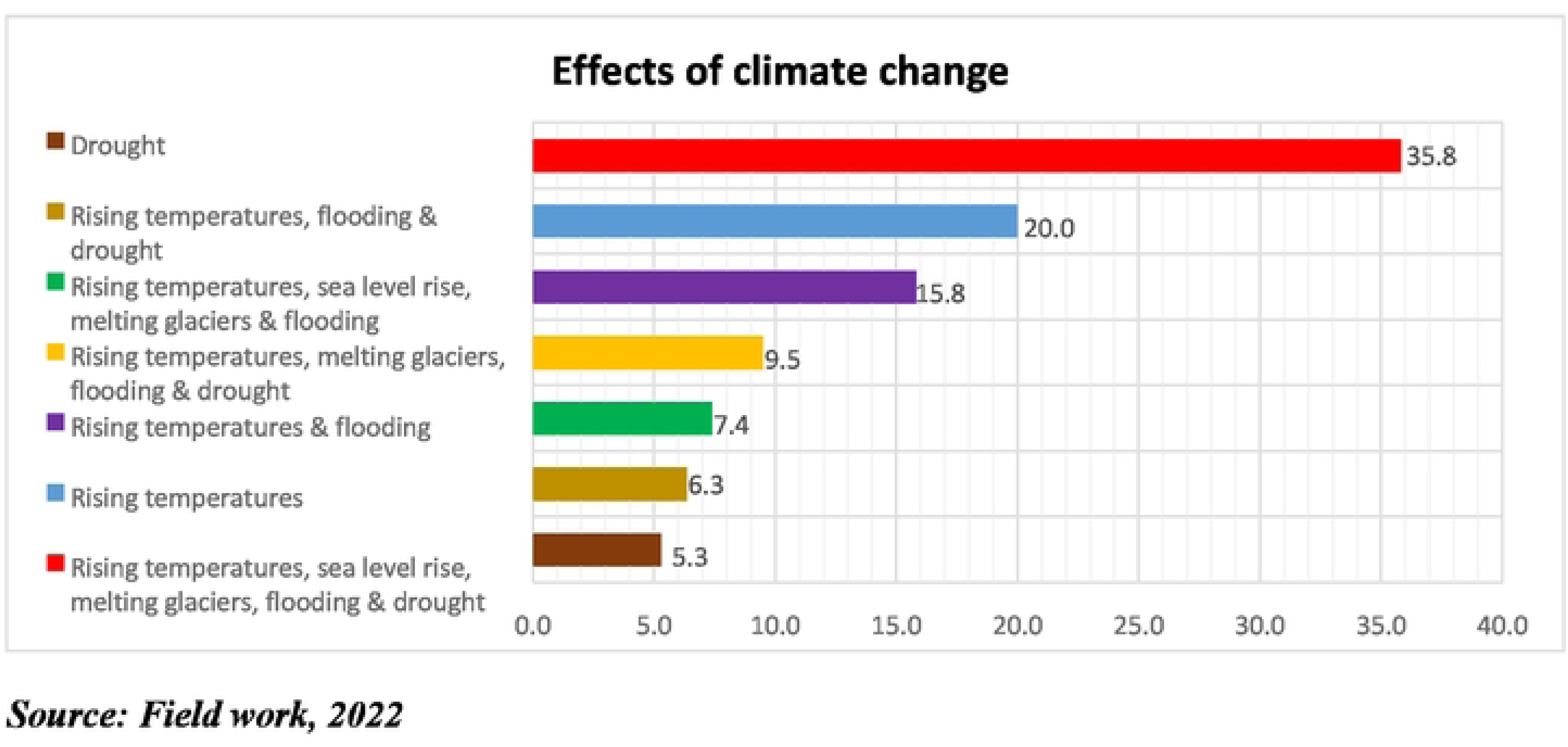
Effects of climate change.

A bivariate analysis was conducted to ascertain the association between the outcome variable (Awareness of the risk of climate change) and various independent variables (background attributes). The results indicate that, gender (P=0.074), age (P=0.443), religious affiliation (P=0.070) and country of study (P=0.331) had no statistical association with respondent’s awareness of the risk of climate change. On the other hand, respondents rank in health service (p<0.05) had a statistical relationship with their awareness of the risk of climate change.

The table 4 above represents health professionals risk perception on climate change. 89% agreed that climate change can lead to death, 8% disagreed and 2% had no idea. Out of those who indicated climate change can lead to death, 79% agreed that climate change is happening worldwide and 8% were unaware of its occurrence worldwide. Majority of the respondents (93%) indicated climate change affects human health while 90% agreed that climate change increases health care cost. Out of 100%, 90% indicated there can be health benefits if climate change is dealt with. Less than half of the respondents (43%) indicated children as the category of people likely to be harmed by the effects of climate change, 18% stated pregnant women, 15% stated the poor and 12% indicated the elderly. 38% of the respondents were moderately concerned about the effects of climate change on health, 27% were extremely concerned, 25% were slightly concerned and 3% were not concerned at all.

**Table 2:** Knowledge on Climate Change (N=400)

| **Attributes** | **Frequency** | **Percentage %** |
| --- | --- | --- |
| **Awareness of climate change** |  |  |
| Yes | 400 | 100 |
| No | 0 | 0 |
| **Human activity is responsible for climate change** |  |  |
| Yes | 348 | 87.0 |
| No | 44 | 11.0 |
| I don't know | 8 | 2.0 |
| **Natural variability is responsible for climate change** |  |  |
| Yes | 298 | 74.5 |
| No | 38 | 9.5 |
| I don't know | 64 | 16 |
| **Extent of human and natural activity on climate change** |  |  |
| Mainly human activity responsible | 52 | 13.0 |
| Mainly natural climate variability responsible | 68 | 17 |
| Solely natural climate variability responsible | 20 | 5.0 |
| Both equally responsible | 260 | 65 |
| **Are climate change awareness campaigns carried out within the Sunyani Municipality** |  |  |
| Yes | 156 | 39.0 |
| No | 152 | 38.0 |
| I don't know | 92 | 23.0 |
| **Which country is most vulnerable to the effects of climate change** |  |  |
| Developed Countries | 30 | 7.5 |
| Developing Countries | 42 | 10.5 |
| Both equally vulnerable | 278 | 69.5 |
| I don't know | 50 | 12.5 |

***Source: Field work, 2022***

**Table 3:** Relationship between Background attributes of respondents and Awareness of the risk of climate change.

| Attributes | **Awareness of the risk of climate change** |  | **P-value** |
| --- | --- | --- | --- |
|  | **Yes; n (%)** | **No; n (%)** |  |
| **Gender** |  |  | .074 |
| Male | 156 (78.7) | 42 (21.3) |  |
| Female | 181 (89.6) | 21 (10.4) |  |

| Age |  |  |  |
| --- | --- | --- | --- |
| 15-24 | 64 (88.9) | 8 (11.1) | .443 |
| 25-34 | 135 (86.5) | 21 (13.5) |  |
| 35-44 | 93 (78.8) | 25 (21.2) |  |
| 45-54 | 29 (87.9) | 4 (12.1) |  |
| 55-60+ | 17 (81) | 4 (19) |  |
| Rank in Health Service |  |  |  |
| Doctor | 17 (57.) | 13 (43) | .020 |
| Nurse | 320 (86.5) | 50 (13.5) |  |
| Religious affiliation |  |  |  |
| Muslim | 80 (100) | 0 (0) | .070 |
| Christian | 224 (77.8) | 64 (22.2) |  |
| African Traditional Religion | 20 (100) | 0 (0) |  |
| No Religion | 12 (0) | 0 (0) |  |
| Country of study |  |  |  |
| Ghana | 333 (84.1) | 63 (15.9) | .331 |
| Outside Ghana | 4 (100) | 0 (0) |  |

***Source: Field work,2022***

**Table 4:**
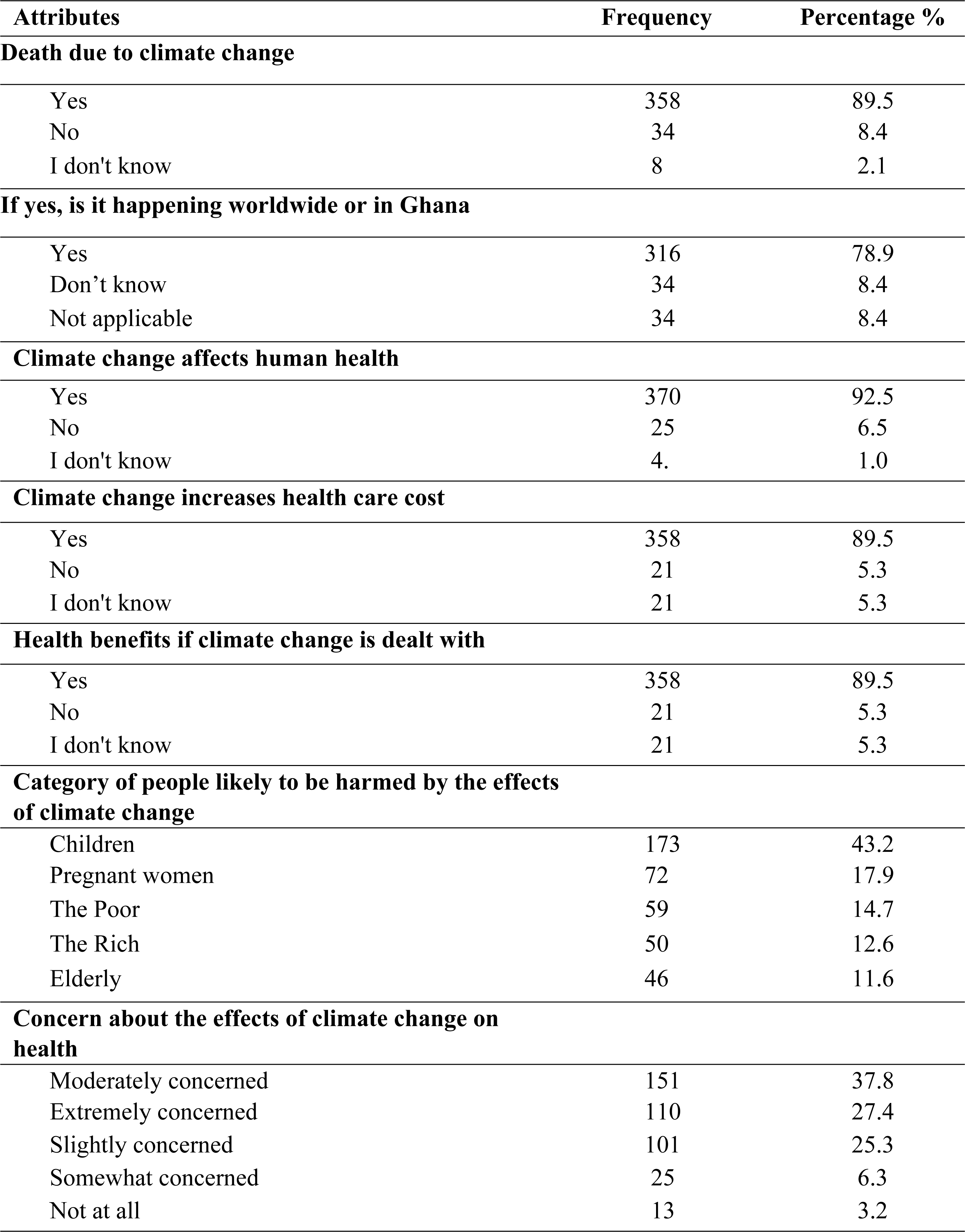
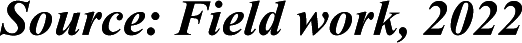
Risk Perception on climate change (N=400)

| **Attributes** | **Frequency** | **Percentage %** |
| --- | --- | --- |
| **Death due to climate change** |  |  |
| Yes | 358 | 89.5 |
| No | 34 | 8.4 |
| I don't know | 8 | 2.1 |
| **If yes, is it happening worldwide or in Ghana** |  |  |
| Yes | 316 | 78.9 |
| Don't know | 34 | 8.4 |
| Not applicable | 34 | 8.4 |
| **Climate change affects human health** |  |  |
| Yes | 370 | 92.5 |
| No | 25 | 6.5 |
| I don't know | 4. | 1.0 |
| **Climate change increases health care cost** |  |  |
| Yes | 358 | 89.5 |
| No | 21 | 5.3 |
| I don't know | 21 | 5.3 |
| **Health benefits if climate change is dealt with** |  |  |
| Yes | 358 | 89.5 |
| No | 21 | 5.3 |
| I don't know | 21 | 5.3 |
| **Category of people likely to be harmed by the effects of climate change** |  |  |
| Children | 173 | 43.2 |
| Pregnant women | 72 | 17.9 |
| The Poor | 59 | 14.7 |
| The Rich | 50 | 12.6 |
| Elderly | 46 | 11.6 |
| **Concern about the effects of climate change on health** |  |  |
| Moderately concerned | 151 | 37.8 |
| Extremely concerned | 110 | 27.4 |
| Slightly concerned | 101 | 25.3 |
| Somewhat concerned | 25 | 6.3 |
| Not at all | 13 | 3.2 |

**Source: Field work, 2022**

A chi-square test between health professionals risk perception on climate change and awareness of the health impact of climate change was conducted. From table 5 above, results indicated a statistical relationship between respondent’s perception that climate change can lead to death (P<0.001) and their awareness of climate change. There was also an association between health benefits if climate change is dealt with (P<0.001) and awareness of the risk of climate change. However, these was no statistical relationship between the health care cost due to climate change (P=0.136), people likely to be harmed by climate change (P=0.425) and respondents awareness of the risk of climate change.

**Table 5:** Relationship between Health professional’s risk perception and Awareness of the health impact of climate change.

| Attributes | Awareness of the risk of climate change |  |  |
| --- | --- | --- | --- |
|  | Yes; n (%) | No; n (%) | P-value |
| Death due to climate change |  |  |  |
| Yes | 337 (94.1) | 21 (5.9) | .001 |
| No | 34 (100) | 0 (0) |  |
| I don't know | 0 (0) | 8 (100) |  |
| Climate change increases health care cost |  |  |  |
| Yes | 337 (94.1) | 21 (5.9) | .136 |
| No | 17 (80) | 4 (20) |  |
| I don't know | 17 (80) | 4 (20) |  |
| Health benefits if climate change is dealt with |  |  |  |
| Yes | 81 (95.3) | 4 (4.7) | .001 |
| No | 5 (100) | 0 (0) |  |
| I don't know | 2 (40) | 3 (60) |  |
| **Category of people likely to be harmed by the effects of climate change** |  |  |  |
| Children | 160 (92.7) | 13 (7.3) | .425 |
| Pregnant women | 64 (88.2) | 8 (11.8) |  |
| The Poor | 55 (92.9) | 4 (7.1) |  |
| The Rich | 46 (91.7) | 4 (8.3) |  |
| Elderly | 46 (100) | 0 (0) |  |
| **Concern about the effects of climate change on health** |  |  |  |
| Moderately concerned | 143 (94.4) | 9 (5.6) | .177 |
| Extremely concerned | 106 (96.6) | 4 (3.8) |  |
| Slightly concerned | 84 (83.3) | 17 (16.7) |  |
| Somewhat concerned | 25 (100) | 0 (0) |  |
| Not at all | 13 (100) | 0 (0) |  |

***Source: Field work,2022***

The table 6 above represents the extent of climate change increasing the likelihood of these diseases. Out of 100%, 34% indicated malaria to be very likely with climate change, 31% stated remotely likely, 24% indicated Dengue fever as very likely, another 24% as unlikely and 15% as likely. Cholera was noted to be very likely by 27%, 14% as possible and 15% as remotely likely. 21% indicated schistosomiasis as very likely to occur while meningococcal meningitis was indicated by 39% as very likely, 26% as likely and 10% as unlikely.

**Table 6:**
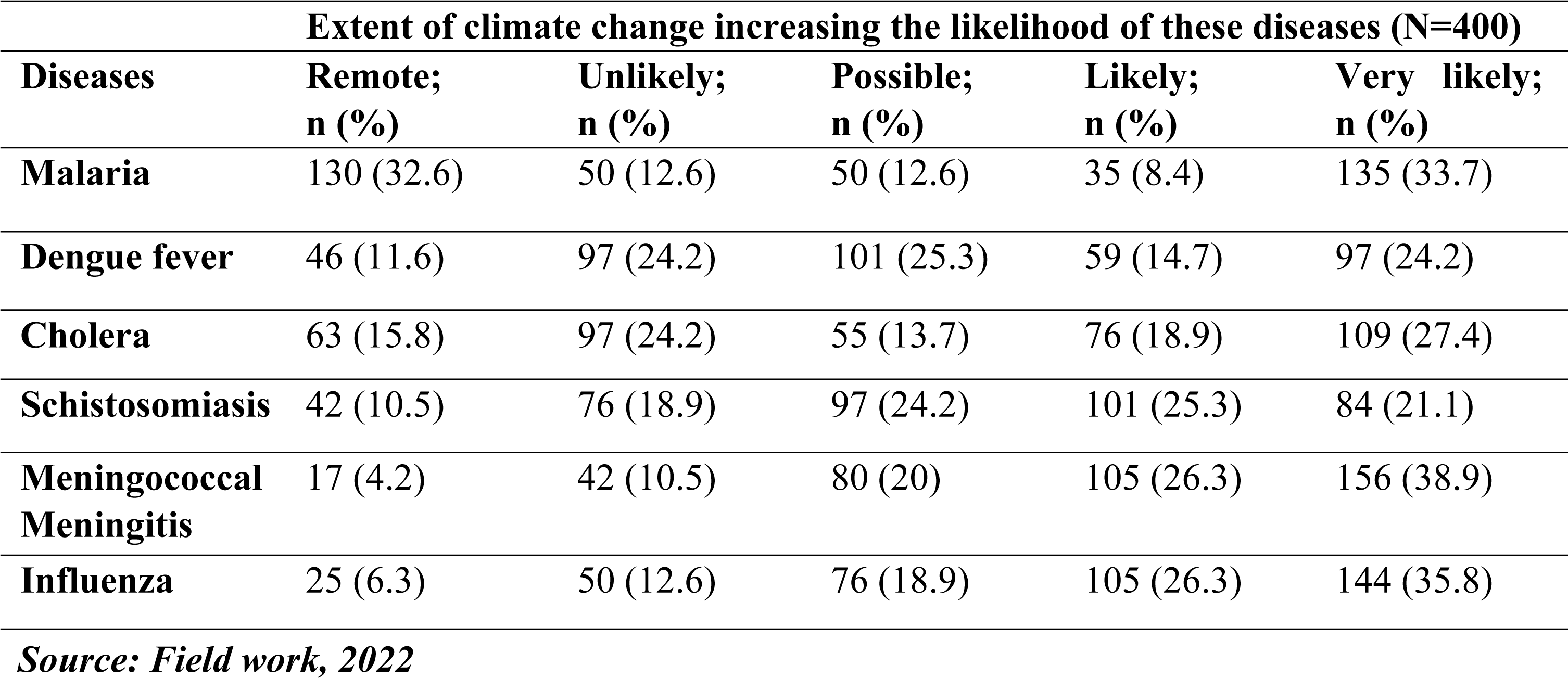
Diseases associated with climate change.

| **Diseases** | **Extent of climate change increasing the likelihood of these diseases (N=400)** |  |  |  |  |
| --- | --- | --- | --- | --- | --- |
|  | **Remote;**<br>**n (%)** | **Unlikely;**<br>**n (%)** | **Possible;**<br>**n (%)** | **Likely;**<br>**n (%)** | **Very likely;**<br>**n (%)** |
| **Malaria** | 130 (32.6) | 50 (12.6) | 50 (12.6) | 35 (8.4) | 135 (33.7) |
| **Dengue fever** | 46 (11.6) | 97 (24.2) | 101 (25.3) | 59 (14.7) | 97 (24.2) |
| **Cholera** | 63 (15.8) | 97 (24.2) | 55 (13.7) | 76 (18.9) | 109 (27.4) |
| **Schistosomiasis** | 42 (10.5) | 76 (18.9) | 97 (24.2) | 101 (25.3) | 84 (21.1) |
| **Meningococcal**<br>**Meningitis** | 17 (4.2) | 42 (10.5) | 80 (20) | 105 (26.3) | 156 (38.9) |
| **Influenza** | 25 (6.3) | 50 (12.6) | 76 (18.9) | 105 (26.3) | 144 (35.8) |

*Source: Field work, 2022*

**Figure 3** displays the problems associated with climate change 18% stated drought as the problem associated with climate change, 15% stated the cancers, 14% noted malaria, 13% skin conditions, another 13% indicated respiratory conditions, 12% stated eye infections and 10% stated flooding. (Find attached)

**Figure 3:**
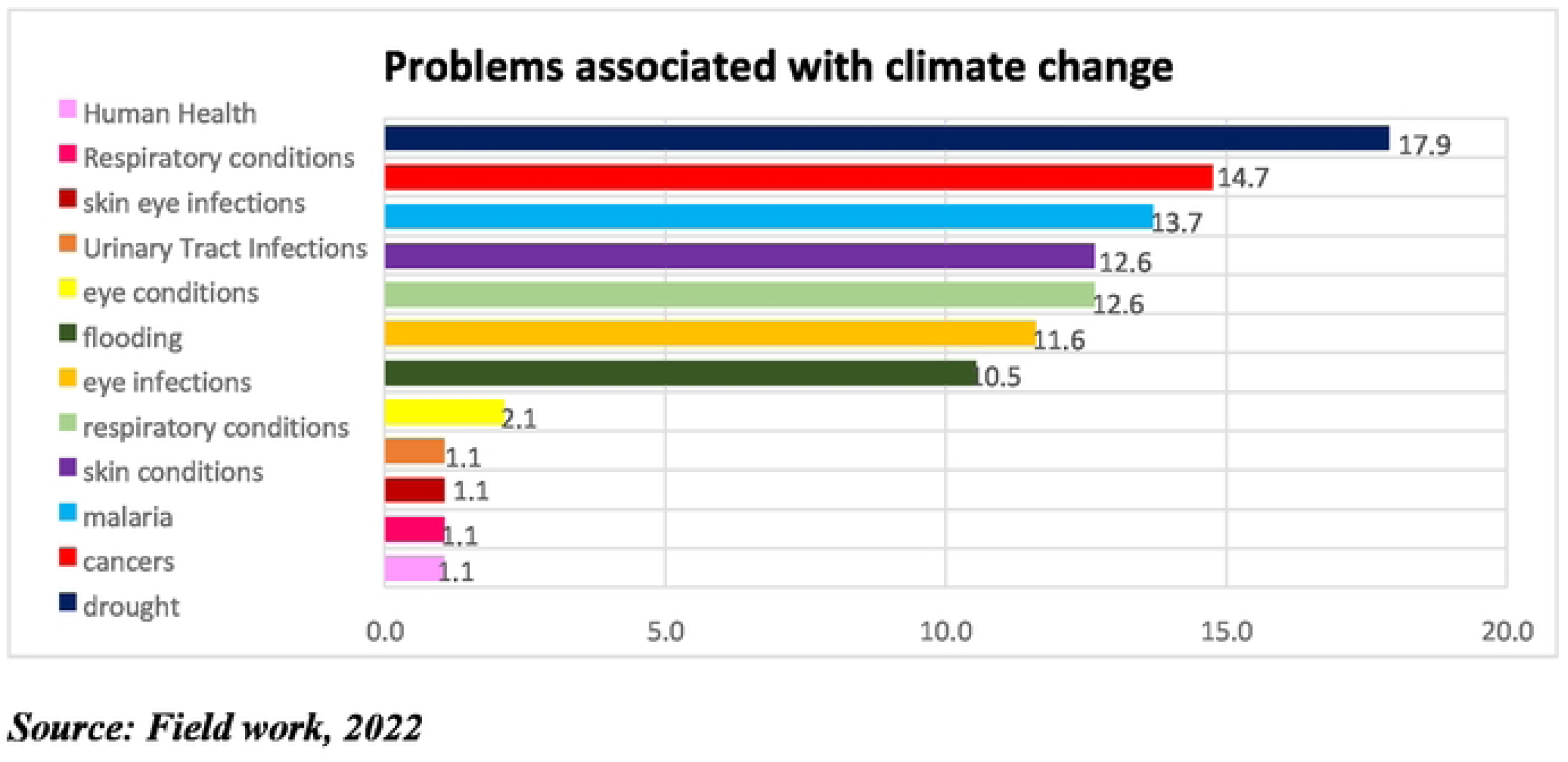
Problems associated with climate change.

A descriptive analysis was conducted, where a Mean score of 2.50 and above indicates a general likelihood to the statement and Mean score below 2.50 indicates a general unlikelihood to the statement. It can be observed that, all six diseases associated with climate change have a Mean score greater than 2.5. It can therefore be concluded that, there was a general likelihood of an increases in malaria (Mean=2.98), Dengue fever (Mean=3.16), Cholera (Mean= 3.18), schistosomiasis (Mean=3.27), Meningococcal meningitis (Mean=3.85) and Influenza (Mean=3.73) due to climate change as indicated by respondents.

Table 8 above shows how health professionals view how climate change can affect Nutrition, Floods, Drought, Mental health and Heat waves. It can be observed that health professionals have a varying understanding of the extent climate change can increase the likelihood of the aforementioned factors.

**Table 7:** Mean and Standard Deviation Score for extent of climate change increasing the likelihood of these diseases.

| **Diseases** | **Mean** | **Standard deviation** |
| --- | --- | --- |
| **Malaria** | 2.98 | 1.701 |
| **Dengue Fever** | 3.16 | 1.347 |
| **Cholera** | 3.18 | 1.466 |
| **Schistosomiasis** | 3.27 | 1.284 |
| **Meningococcal meningitis** | 3.85 | 1.176 |
| **Influenza** | 3.73 | 1.250 |

*Source: Field work, 2022*

**Table 8:** Risk of climate change.

| **Factors** | **Extent of climate change increasing the likelihood of these factors (N=400)** |  |  |  |  |
| --- | --- | --- | --- | --- | --- |
|  | **Remote;**<br>**n (%)** | **Unlikely;**<br>**n (%)** | **Possible;**<br>**n (%)** | **Likely;**<br>**n (%)** | **Very likely;**<br>**n (%)** |
| **Nutrition** | 97 (24.2) | 38 (9.5) | 59 (14.7) | 63 (15.8) | 143 (35.8) |
| **Floods** | 42 (10.5) | 63 (15.8) | 59 (14.7) | 34 (8.4) | 202 (50.5) |
| **Drought** | 67 (16.8) | 42 (10.5) | 84 (21.1) | 63 (15.8) | 143 (35.8) |
| **Mental health** | 97 (24.2) | 59 (14.7) | 97 (24.2) | 54 (13.7) | 93 (23.2) |
| **Heat waves** | 46 (11.6) | 63 (15.8) | 63 (15.8) | 72 (17.9) | 156 (38.9) |

*Source: Field work, 2022*

A descriptive analysis was conducted, where a Mean score of 2.50 and above indicates a general likelihood to the statement and Mean score below 2.50 indicates a general unlikelihood to the statement. It can be observed from table 9 above that all five factors have a Mean Score more than 2.50 and hence there is a general likelihood of nutrition (Mean=3.29), floods (Mean=3.73), drought (Mean=3.43), mental health (Mean=2.97) and heat waves (Mean=3.57) being increased by climate change as indicated by respondents.

**Table 9:** Mean and Standard Deviation Score for extent of climate change increasing the likelihood of these factors.

| **Factors** | **Mean** | **Standard deviation** |
| --- | --- | --- |
| **Nutrition** | 3.29 | 1.610 |
| **Floods** | 3.73 | 1.476 |
| **Drought** | 3.43 | 1.485 |
| **Mental health** | 2.97 | 1.484 |
| **Heat waves** | 3.57 | 1.434 |

*Source: Field work, 2022*

On the actions towards climate change mitigation, 35% indicated buying and using energy light bulbs and appliances as very likely to mitigate climate change. The use of less air conditioning during dry season was indicated by 28% as very likely, 23% as likely and 30% as unlikely. Purchase and use of solar panels was stated as very likely by 31%, 14% as unlikely and 29% as remotely likely. Car pool at least a couple of days per week was stated by 30% as very likely and 32% as remotely likely. The use of public transport even if an individual owns a vehicle was noted by 16% as very likely, 19% as unlikely and 30% as remotely unlikely. Finally, walking and cycling instead of using cars always was indicated by 30% as very likely in mitigating climate change, 10% as unlikely and 23% as remotely unlikely.

A descriptive analysis was conducted, where a Mean score of 2.50 and above indicates a general likelihood to the statement and Mean score below 2.50 indicates a general unlikelihood to the statement. It can be observed that all actions towards climate change mitigation have a Mean Score greater than 2.50. Therefore, it can be concluded that, there is a greater likelihood of mitigating climate change through these actions as indicated by the respondents.

Certain factors were identified to be beneficial towards climate change mitigation efforts, and table 12 above shows the degree with which health professionals see those factors as beneficial to the climate change mitigation efforts.

**Table 10:** Actions towards climate change mitigation.

| **Willingness to act towards climate change (N=400)** |  |  |  |  |  |
| --- | --- | --- | --- | --- | --- |
| **Actions** | **Remote;**<br>**n (%)** | **Unlikely;**<br>**n (%)** | **Possible;**<br>**n (%)** | **Likely;**<br>**n (%)** | **Very likely;**<br>**n (%)** |
| **Buy and use energy light bulbs and appliances** | 93 (23.2) | 72 (17.9) | 38 (9.5) | 59 (14.7) | 139 (34.7) |
| **Use less air conditioning during dry season** | 38 (9.5) | 118 (29.5) | 38 (9.5) | 93 (23.2) | 114 (28.4) |
| **Purchase and use solar panels** | 118 (29.5) | 59 (14.7) | 34 (8.4) | 63 (15.8) | 122 (30.5) |
| **Car pool at least a couple of days per week** | 126 (31.6) | 17 (4.2) | 72 (17.9) | 63 (15.8) | 122 (30.5) |
| **Use public transport even if you own a vehicle** | 122 (30.5) | 76 (18.9) | 46 (11.6) | 93 (23.2) | 63 (15.8) |
| **Walking and cycling instead of using cars always** | 93 (23.2) | 38 (9.5) | 59 (14.7) | 88 (22.1) | 122 (30.5) |

*Source: Field work, 2022*

**Table 11:**
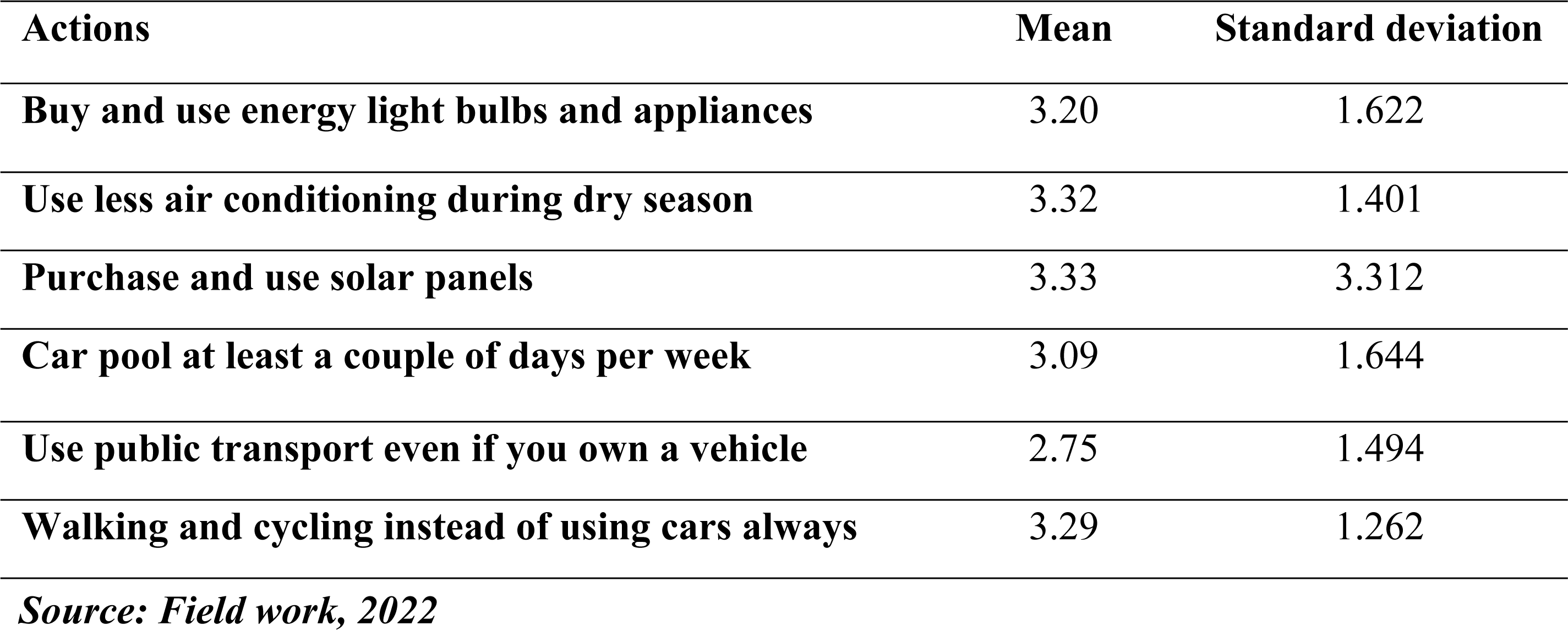
Mean and Standard Deviation Score for actions towards climate change (N=400)

**Table 12:**
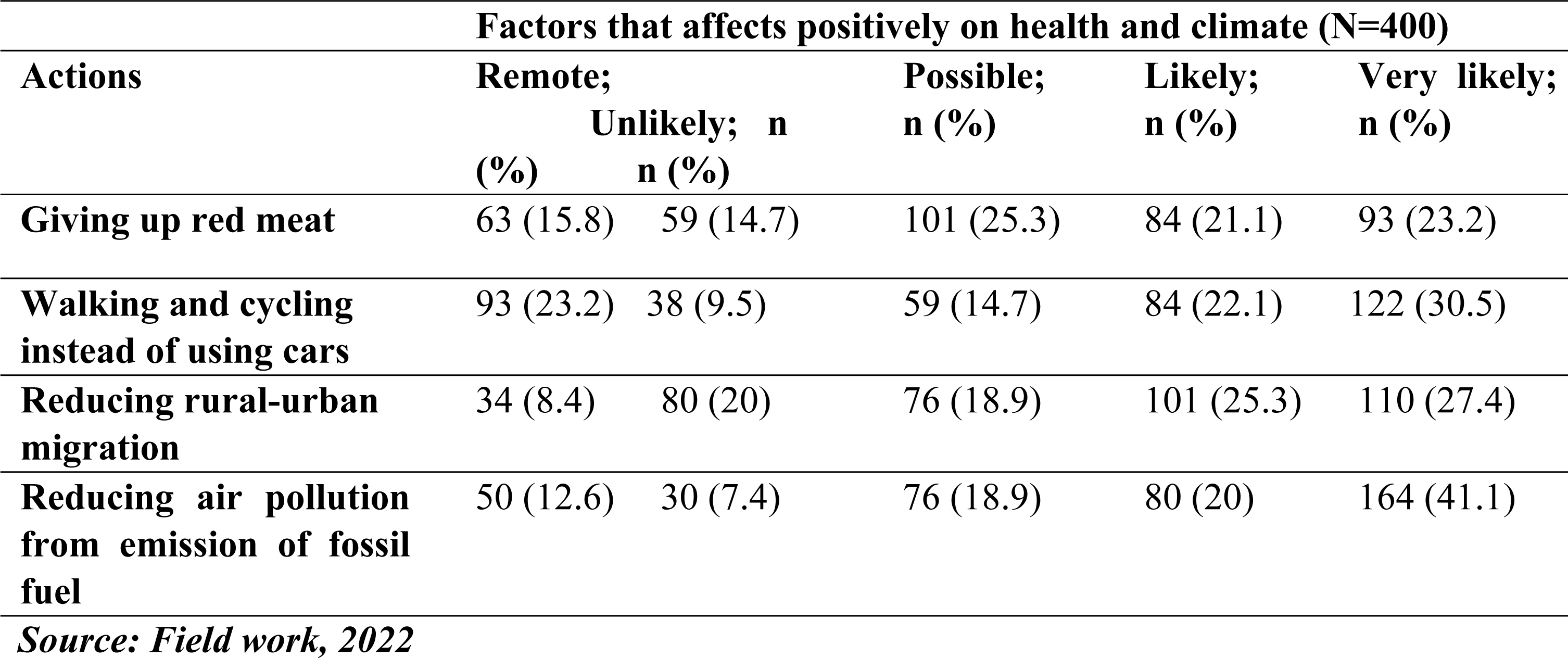
Co-benefits of climate change mitigation.

| **Actions** | **Factors that affects positively on health and climate (N=400)** |  |  |  |  |
| --- | --- | --- | --- | --- | --- |
|  | **Remote;**<br>**(%)** | **Unlikely; n**<br>**(%)** | **Possible;**<br>**n (%)** | **Likely;**<br>**n (%)** | **Very likely;**<br>**n (%)** |
| **Giving up red meat** | 63 (15.8) | 59 (14.7) | 101 (25.3) | 84 (21.1) | 93 (23.2) |
| **Walking and cycling instead of using cars** | 93 (23.2) | 38 (9.5) | 59 (14.7) | 84 (22.1) | 122 (30.5) |
| **Reducing rural-urban migration** | 34 (8.4) | 80 (20) | 76 (18.9) | 101 (25.3) | 110 (27.4) |
| **Reducing air pollution from emission of fossil fuel** | 50 (12.6) | 30 (7.4) | 76 (18.9) | 80 (20) | 164 (41.1) |

*Source: Field work, 2022*

A descriptive analysis was conducted, where a Mean score of 2.50 and above indicates a general likelihood to the statement and Mean score below 2.50 indicates a general unlikelihood to the statement. It can be observed from table 13 that all actions that positively affects health and climate have a mean score greater than 2.50. it can be concluded that, the likelihood of these actions impacting positively on climate change mitigation is very high.

**Table 13:** Mean and Standard Deviation Score for factors that positively affects health and climate (N=400)

| **Actions** | **Mean** | **Standard deviation** |
| --- | --- | --- |
| **Giving up red meat** | 3.21 | 1.375 |
| **Walking and cycling instead of using cars** | 3.27 | 1.554 |
| **Reducing rural-urban migration** | 3.43 | 1.310 |
| **Reducing air pollution from emission of fossil fuel** | 3.69 | 1.400 |

*Source: Field work, 2022*

From the table 14 above, majority of the respondents (85%) agreed to environmentally sustainable and energy practices at all health facilities as one of the policies to mitigate climate change, 94% indicated encouraging health professionals to train in climate change and its effects and 65% agreed to the incorporation of climate change related course work into nursing/medical school.

**Table 14:**
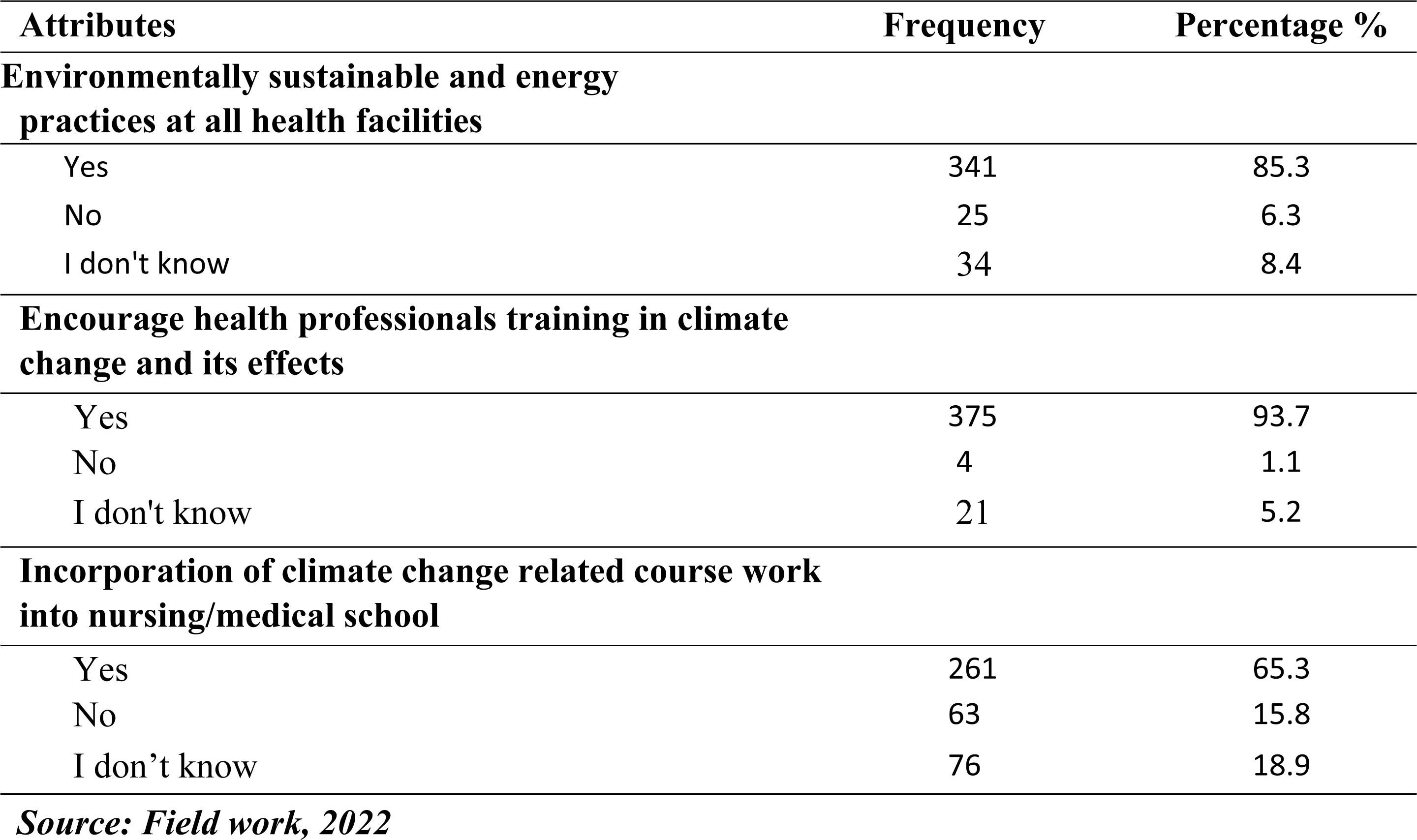
Policy and practice for climate change (N=400)

## Discussion

### Health Professional’s Knowledge on Climate Change

Since climate change poses and existential threat to humans, public knowledge and awareness are key ingredients that will aid in addressing the canker. Sound knowledge of climate change is believed to be a viable antidote necessary to address the problem because knowledge shapes people’s behavior to put up attitudes which will inure to the benefit of the environment and thus eschew attitudes which contribute to global warming. ^2,3^

From this current study, all respondents (100%) were fully aware about the concept of climate change. This corroborates the revelation by ^9^ in a study conducted on the general perceptions of people towards the concept of climate change in Africa, where authors observed strict evidence of a general awareness of the concept of climate change across the various African communities they studied.

With respect to the extent of human and natural activity on climate change, more than half (65%) of the respondents indicated that both human and natural activity are responsible for climate change, this finding affirms the observations made by ^8^ who opined that the changes in global climate occasioned by both internal changes within the climate ecosystem and external variabilities including both human-induced and naturally induced factors have evidently shown that weather and climate are always. Majority of the respondents (87%) agreed that human activity is responsible for climate change, this affirms the evidence given by ^23^ that humans are changing earth’s climate causing the atmosphere and oceans to be warmed.

Another majority of 75% of respondents noted natural variability as responsible for climate change, this current observation by the researchers concerning health professionals in the Sunyani Municipality means that, indeed they have knowledge about the fact that natural variability is responsible for climate change, however they don’t understand the scientific basis for this based on the verbal questions the researchers posed to the respondents concerning it. This finding affirms the observation made by ^19^ where they opined that famers including many others like them have the knowledge about climate change, however there is the need for more education on the scientific aspects and causes of climate change.

Majority of respondents (70%) emphasized that both developed and developing countries are equally vulnerable to the effects of climate change, this finding corroborates the observations made by ^6^ who opined that history records widespread disasters, famines and disease outbreaks triggered by droughts and floods which are exerting worst effects in poor countries but even the richest industrial societies are not immuned. This finding however means that, the effects of climate change is no respecter of country, irrespective of whether that country is economically advanced or not. Hence, the phenomenon of climate change should be given equal attention by all countries across the globe.

### Health Professional’s risk perception on climate change

The mental and social picture towards climate change is largely influenced by the perception of individuals. However, it is important to add that catalysts such as ideological orientations of individuals, socio-economic and socio-demographic characteristics of individuals all influence the perception and the mental picture individuals carve about climate change. ^10^ Many definitions have been given to explain risk but in all of it, the underlying factor is that, there is the likelihood of an individual experiencing the effect of danger.^3^ It also defined as the probability of an adverse effect and the magnitude of its consequence. All risk concepts have one element in common; a distinction between reality and possibility.^3^

From this current study, the results indicate that, gender (P=0.074), age (P=0.443), religious affiliation (P=0.070) and country of study (P=0.331) had no statistical association with respondent’s awareness of the risk of climate change. This is due to the fact that, a P value greater than 0.05 is an indication of no statistical relationship between the two variables. This however means that, the gender, age, religious affiliation and country of study of health professionals within the Sunyani Municipality has nothing to do with their perception of risk on climate change. On the other hand, respondents rank in health service (p<0.05) had a statistical relationship with their awareness of the risk of climate change. This means a health professional’s rank in health service increases their perfection of risk of climate change, i.e. the higher the rank in health service, the greater the awareness and perception of risk of climate change.

The findings from this current study further indicates that, a majority of 89% of health professionals within the Sunyani municipality had the perception of risk that climate change can lead to death. This finding affirms the position of ^17^ who opined that, in 1998 the severe flooding that hit West Bengal was accompanied by cholera outbreak which triggered 16,590 cases of the diarrheal disease and further led to 276 deaths with a 1.7% case fatality rate. The study further found a statistical relationship between respondent’s perception that climate change can lead to death (P<0.001) and their awareness on the health impact of climate change. This means that perception of death as a result of climate change makes health professionals within the Sunyani municipality aware of the negative health impacts posed by climate change. Majority of the respondents (93%) had a risk perception that climate change affects human health, this however means that 93% of health professionals within the Sunyani municipality had a perception that climate change can affect human health. This finding corroborates the observation made by ^24^ where they stated that, climate change has a higher propensity to affect human health through indirect routes especially through ecological and biological processes which drives communicable disease transmission.

Another majority of 90% of respondents had the perception that climate change increases health care cost. This is an indication of the fact that 90% of health professionals within the Sunyani municipality had the perception that climate change has the potentiality of increasing health care cost. This finding affirms the position of ^26^ where they observed that, in the WHO Europe region, there is a burden of climate change, including premature deaths and morbidity and is estimated to be equivalent to around $1.6 trillion in 2010 represents 10% or more of GDP in almost half of the countries covered.

It was further observed that 90% of respondents had the perception that there can be health benefits if climate change is dealt with. Less than half of the respondents (43%) had the perception that children are the category of people likely to be harmed by the effects of climate change, 18% stated pregnant women, 15% stated the poor and 12% indicated the elderly. 38% of the respondents were moderately concerned about the effects of climate change on the health of people within the Sunyani municipality, 27% were extremely concerned, 25% were slightly concerned and 3% were not concerned at all.

On the effects of climate change, 36% of respondents believed rising temperature, sea level rise, melting glaciers and drought are some of the effects of climate change. This finding corroborates the observation made by ^28^ who opined that, the atmosphere and oceans have warmed, accompanied by sea-level rise, a strong decline in Arctic sea ice, and other climate-related changes.

Again, 20% of respondents indicated rising temperature as an effect of climate change. Another 16% of health professionals noted rising temperature and flooding as the effects of climate change. The remaining 10% of the respondents indicated rising temperature, melting glaciers, flooding and drought as the effects of climate change and this finding from this current study affirms the position of ^17^ who observed that while drought which is a natural phenomenon caused by changes in weather patterns due to reduced or less rainfall than usual reduces the water available for washing and sanitation and also tends to increase the risk of disease.

It was further found that, there was a general likelihood of an increases in malaria (Mean=2.98), Dengue fever (Mean=3.16), Cholera (Mean= 3.18), schistosomiasis (Mean=3.27), Meningococcal meningitis (Mean=3.85) and Influenza (Mean=3.73) due to climate change as indicated by respondents who are health professionals. The above findings simply mean that climate change has a higher probability of causing the above-named diseases (Dengue fever, Cholera, Schistosomiasis, Meningococcal meningitis and Influenza) as indicated by health professionals in the study. These findings corroborate what similar studies found previously. ^7,8^

It was further observed there is a general likelihood of nutrition (Mean=3.29), floods (Mean=3.73), drought (Mean=3.43), mental health (Mean=2.97) and heat waves (Mean=3.57) being increased by climate change as indicated by respondents. These findings simply mean that, climate change has a high probability to increase nutritional challenges (disorders), increase floods, increase droughts, increase mental health challenges (disorders) as well as increase heat waves as indicated by respondents who are health professionals in the study. These findings are in tandem with similar observations made by other researchers. ^5,6^

### Co-benefits of climate change mitigation

The immense benefits of climate change mitigation cannot be overemphasized as it is the lead pathway towards the newly envisioned renewable energy world.^26^ There are many activities that are capable of reducing Green House Gas (GHG) emissions and its benefits goes beyond contributing to climate change mitigation alone. From the UNECE, 2016 report it was stated that, the drastic reduction of the emissions of fossil fuels through pollution of the air are the most visible co-benefit of climate change mitigation and wields a positive health and environmental benefits to people. Furthermore, areas like heightened sustainability of ecosystems, efficiency use of natural resources and economic security are the other benefits climate change mitigation can bring to human populations.

From this current study, in order to ascertain the potency of some actions towards climate change mitigation, it was observed that all actions towards climate change mitigation were capable of ameliorating the menace. These actions included: Buying and use energy light bulbs and appliances (Mean=3.20), Using less air conditioning during dry season (Mean=3.32), Purchasing and using solar panels (Mean=3.33), Carpooling at least a couple of days per week (Mean=3.09), Using public transport even if you own a vehicle (Mean=2.75), Walking and cycling instead of using fuel-based cars always (Mean=3.29). These findings agree with earlier observations found in literature.^26^

This current study further tried to establish the potency of some factors that impact positively on health and climate It can be observed that all these actions positively affect health and climate. Such actions included: Giving up red meat (Mean=3.21), Walking and cycling instead of using cars (Mean=3.27), Reducing rural-urban migration (Mean=3.46), Reducing air pollution from emission of fossil fuel (Mean=3.63). This finding affirms the position of UNECE (2016) where its report stated that, the drastic reduction in fossil fuel emission through pollution of the air and the accompanying environmental benefits are the most visible co-benefit of climate change mitigation. It can therefore be concluded that, the likelihood of these actions impacting positively on climate change and health is very high as indicated by health professionals in the study.^26^

On policy and practice for climate change mitigation, majority of the respondents (85%) agreed to environmentally sustainable and energy practices at all health facilities as one of the policies to mitigate climate change. It is worth noting the fact that climate change mitigation largely involves the reduction in human emissions of greenhouse gases into the atmosphere. Mitigation can further be achieved through the increment carbon sinks in the environment, such as through reforestation at the various health facilities. Climate change mitigation can also be achieved through the adoption of low-carbon energy use (such as renewable and nuclear energy) thereby phasing out the use of fossil fuels. ^29, 30^

Another majority of 94% of respondents indicated encouraging health professionals to train in climate change and its effects and an additional 65% of respondents agreed to the incorporation of climate change related course work into nursing/medical school curricula as some policies to mitigate climate change. These findings corroborate earlier observations made by other researchers. ^29^

## Summary of Findings

### Health Professional’s Knowledge on Climate Change

This segment of the study sought to assess the knowledge level of health professionals on climate change. It came to light that:

- Health professionals are fully aware of the concept of climate change and therefore have some form of knowledge about it.
- Health professionals do not have adequate knowledge pertaining to the scientific aspects of climate change.

### Health Professional’s risk perception on climate change

This section of the study sought to examine the health professional’s perception of risk in respect of climate change. It came to light that:

- There was a statistical relationship between respondent’s perception that climate change can lead to death (P<0.001) and their awareness of the risk of climate change impact on health.
- There was also a statistical association between health benefits if climate change is dealt with (P<0.001) and awareness of the risk of climate change impact on health.
- There was no statistical association between health care cost due to climate change (P=0.136), people likely to be harmed by climate change (P=0.425) and respondents awareness of the risk of climate change impact on health.
- There was a general likelihood of an increase in malaria (Mean=2.98), Dengue fever (Mean=3.16), Cholera (Mean= 3.18), schistomiasis (Mean=3.27), Meningococcal meningitis (Mean=3.85) and Influenza (Mean=3.73) due to climate change.
- There was a general likelihood of nutrition (Mean=3.29), floods (Mean=3.73), drought (Mean=3.43), mental health (Mean=2.97) and heat waves (Mean=3.57) being increased due to climate change.

### Co-benefits of climate change mitigation

This portion of the study sought to verify the co-benefits of climate change mitigation. The study found that:

- There is a greater likelihood of mitigating climate change through these actions: Buy and use energy light bulbs and appliances (Mean=3.20), Use less air conditioning during dry season (Mean=3.32), Purchase and use solar panels (Mean=3.33), Car pool at least a couple of days per week (Mean=3.09), Use public transport even if you own a vehicle (Mean=2.75), Walking and cycling instead of using cars always (Mean=3.29).
- These actions positively affect health and climate and they include: Giving up red meat (Mean=3.21), Walking and cycling instead of using cars (Mean=3.27), Reducing ruralurban migration (Mean=3.46), Reducing air pollution from emission of fossil fuel (Mean=3.63).
- Majority of the respondents (85%) agreed to environmentally sustainable and energy practices at all health facilities as one of the policies to mitigate climate change.
- 94% of respondents indicated encouraging health professionals to train in climate change and its effects as another policy to mitigate climate change.
- 65% of respondents agreed to the incorporation of climate change related course work into nursing/medical school curricula as a policy to mitigate climate change.

## Conclusions

Health professionals are fully aware about climate change but lack a thorough understanding of the scientific aspects of climate change. The general risk perception of health professionals towards climate change impact on health was high. Climate change mitigation is beneficial to human populations.

## Recommendations

Based on the findings and conclusions from the study (investigating health worker understanding and risk perception of climate change to health in the Sunyani Municipality, Ghana), the following recommendations are suggested:

- The Ministry of Health and the Ghana Health Service should intensify climate change awareness campaigns across the length and breadth of the country in order to create more awareness about the phenomenon.
- Health management and authorities at the various health facilities in the country should provide intense information and education to health professionals concerning climate so as to enable them understand the scientific dimensions of climate change.
- Based on the grievous effects of climate change, the Government of Ghana should make it a national policy to include into the laws of Ghana, a constitutional provision forbidding people from excessive emissions of greenhouse gases into the environment with corresponding prescribed punishment for such acts.
- The Government of Ghana should again include into the factories, offices and shop Acts, a clause which correctly spells out acceptable threshold of chemical emissions into the environment which all factories and companies in the country have to comply with.
- The Ministry of Health and the Ghana Health Service should design a policy document that would ensure that all health institutions in the country subscribe to actions to mitigate climate change through an environmentally sustainable and energy practices, such reforestation at the various health facilities and also phasing out fossil fuels by switching to low-carbon energy sources such as renewable energy e.g. solar.

### Avenue for further research

- Another scientifically oriented research is recommended to look in-depth into the relationship between climate change and non-communicable diseases and suggest recommendations that are medically feasible.

## Authors’ contribution

**AJB** and AGM conceptualized the overall study with its goals and aims. BOA retrieved the requisite data from databases. RDC and HG played a supportive role in data consolidation. SA wrote the study background and played a supportive role in data analysis. DA played a role in designing the methodology. YBN played a vital role in designing the methodology for the study. JD played a role in writing discussion of the study. LA supported in the writing, review and editing of the of the manuscript. BOA, AOT and LA did the data analysis for the study.

All authors contributed significantly to the critical revision and approved the final version before the onward submission.

## Data Availability

Data is always available

## Acknowledgement

We express our heartfelt gratitude to the health professionals specifically doctors and nurses at the Regional Hospital, SDA Hospital and Sunyani Municipal Hospital for their willingness to partake in the study.

